# Smartphone video–based knee extension moments during chair rise relate to MRI measures of muscle function

**DOI:** 10.64898/2026.03.08.26347617

**Authors:** R. Daniel Magruder, Mary Hall, Yael Vainberg, Jessica L. Asay, Feliks Kogan, Jennifer L. Hicks, Garry E. Gold, Scott L. Delp, Scott D. Uhlrich, Valentina Mazzoli

## Abstract

**Background:** Preserving muscle function is essential for maintaining independence during aging, but muscle force-generating capacity is not commonly measured clinically due to a lack of accessible, sensitive tools. Magnetic resonance imaging (MRI) provides gold-standard measures of muscle volume and microstructure, which reflect force-generating capacity, while dynamometry quantifies peak joint moments during voluntary contraction. Both modalities are time-consuming and costly, so clinical and large-scale studies often rely on low-fidelity measures such as the time to complete the five times sit-to-stand test (5xSTS). OpenCap, a tool for quantifying musculoskeletal dynamics from smartphone videos, may provide an accessible and more informative approach to assessing muscle function. We evaluated whether OpenCap-derived knee extension moments during chair rise relate to MRI-based measures of quadriceps muscle volume and microstructure, using dynamometry as a comparator.

**Methods:** Nineteen healthy adults of various ages (63.2% female, 57.8 ± 15.4 y, 30–78 y) underwent quadriceps MRI, dynamometry, and 5xSTS time with concurrent OpenCap data collection. Using MRI, we computed quadriceps volume and radial diffusivity (a measure related to fiber size). We standardized these features and summed to create a composite MRI score, reflecting muscle quantity and quality. We estimated peak knee extension moment using OpenCap during chair rise and via both isometric and isokinetic dynamometry. We compared OpenCap kinematics (torso angle) and dynamics (knee moment), 5xSTS time, and dynamometry to MRI measures of muscle function using linear regression; false discovery rate was controlled using the Benjamini-Hochberg procedure.

**Results:** The OpenCap-derived knee extension moment was associated with quadriceps muscle volume (r=0.63, p=0.014) and radial diffusivity (r=0.61, p=0.016). Peak knee extension moments measured by both isometric and isokinetic dynamometry were correlated with muscle volume (r=0.66–0.75, p=0.002–0.009), but not with radial diffusivity (r=0.04–0.52, p=0.054–0.91). Both OpenCap and isokinetic dynamometry showed their strongest associations with the composite MRI score (r=0.77, p=0.002 and r=0.73, p=0.002, respectively). 5xSTS time and a kinematic feature (torso angle) were not associated with any MRI-derived measures (r=−0.16–0.35, p=0.22–0.97).

**Conclusions:** Smartphone video–based joint moments associate with muscle size and microstructure, unlike time or kinematic features. OpenCap offers a scalable assessment of muscle force-generating capacity that can be conducted rapidly without specialized equipment, enabling higher-fidelity assessments of muscle function in the clinic and in large-scale studies where imaging and dynamometry are impractical.

## Introduction

Maintaining muscle function is critical for sustaining mobility, balance, and independence as we age [1], [2]. Sarcopenia, a degenerative condition characterized by muscle wasting, accelerates age-related decline of muscle strength and contributes to frailty [3]. Early diagnosis is critical but challenging because of its overlap with normal aging and comorbidities [4]. Current clinical screening tools (e.g., grip strength, timed function tests) are practical, but lack specificity [5]. There remains a need for sensitive, scalable markers that can detect deficits in muscle function before irreversible functional decline, enabling early intervention [6].

Magnetic resonance imaging (MRI) captures measures of muscle volume and quality that are associated with force-generating capacity. Muscle volume correlates strongly with force production [7], predicts fall risk [8], and is related to the presence of sarcopenia [9]. Yet muscle force declines faster with age than muscle volume [10], underscoring the importance of muscle quality in force production [11], [12], [13]. Quantitative MRI techniques are sensitive to muscle microstructure and composition, collectively termed muscle quality [14]. For example, diffusion tensor imaging can track water diffusion along and perpendicular to muscle fibers. Higher diffusivity perpendicular to the fibers—known as radial diffusivity—associates with both increased fiber size and a greater proportion of type II fibers [15], [16], features that are associated with force and power production [17]. However, despite its utility, MRI is costly and inaccessible [9], making it impractical for routine screening of muscle function.

Dynamometry measures maximal voluntary joint moments and is used to assess muscle force-generating capacity under static (isometric) and constant angular-velocity (isokinetic) conditions. These have been associated with MRI measures of muscle size and quality [13], [18] and are regarded as the gold standard for assessing strength in sarcopenia [19]. Isokinetic dynamometers are large, costly, require trained operators, and have limited clinical scalability [20]. Hand-held dynamometers address portability [21] but still require trained operators and lack accuracy, especially for strong muscle groups such as the quadriceps [22], [23]. The quadriceps are of particular relevance to sarcopenic and frail populations due to their contribution to fall risk and performing activities of daily living [5], [24], [25].

High-fidelity assessments of muscle function, including MRI and isokinetic dynamometry, are often inaccessible and therefore not routinely conducted clinically. Lower fidelity timed function tests, like the five times sit-to-stand (5xSTS) test, are commonly used in aging and sarcopenia research instead [26]. Rising from a chair as quickly as possible requires large quadriceps forces, and the time to complete the 5xSTS worsens with age [27]. There are several ways to rise from a chair to compensate for quadriceps weakness [28], [29], but by only measuring the time to completion, the 5xSTS does not account for these compensations. This makes the 5xSTS time relatively insensitive to declines in quadriceps function that occur before performance time is affected. Detecting these early changes is essential to enable intervention before irreversible loss of physical function; thus, more accessible, sensitive assessments of lower-extremity muscle function are needed.

Advancements in mobile sensing have democratized access to in-depth biomechanical assessment in the clinic [30], [31], [32]. For example, we developed OpenCap, an open-source platform which uses two or more smartphone videos to quantify kinematics (e.g., joint angles) and kinetics (e.g., joint moments) during functional activities. An OpenCap assessment of the sit-to-stand test can be conducted in the clinic in five minutes [33]. Knee moments during chair rise from lab-based motion capture have been shown to relate to isokinetic measures of muscle strength [34]. Additionally, OpenCap can detect differences in the knee extension moment that arise from common compensatory strategies during chair rise, such as increased trunk lean and angular velocity [28], [29], [35], [36]. Thus, using OpenCap to assess knee moments during the 5xSTS is a promising candidate for an accessible and sensitive assessment of lower-extremity muscle function. However, it remains unknown whether it is sensitive to differences in MRI-derived measures of muscle force-generating capacity across a wide age range.

The purpose of this study was to determine whether OpenCap smartphone video–based measures of quadriceps force (the knee extension moment) could serve as an accessible proxy for MRI measures of quadriceps force-generating capacity. We hypothesized that in a cohort of healthy participants of various ages, the OpenCap-derived peak knee extension moment would correlate with quadriceps volume and radial diffusivity from MRI (Fig. 1). To contextualize these associations with other clinically accessible functional measures, we also related dynamometry, 5xSTS time, and OpenCap kinematics to the MRI metrics. As an additional exploratory step, we combined volume (quantity) and radial diffusivity (quality) into a composite score to test whether functional assessments relate better to integrated measures of muscle force-generating capacity.

**Fig. 1:**
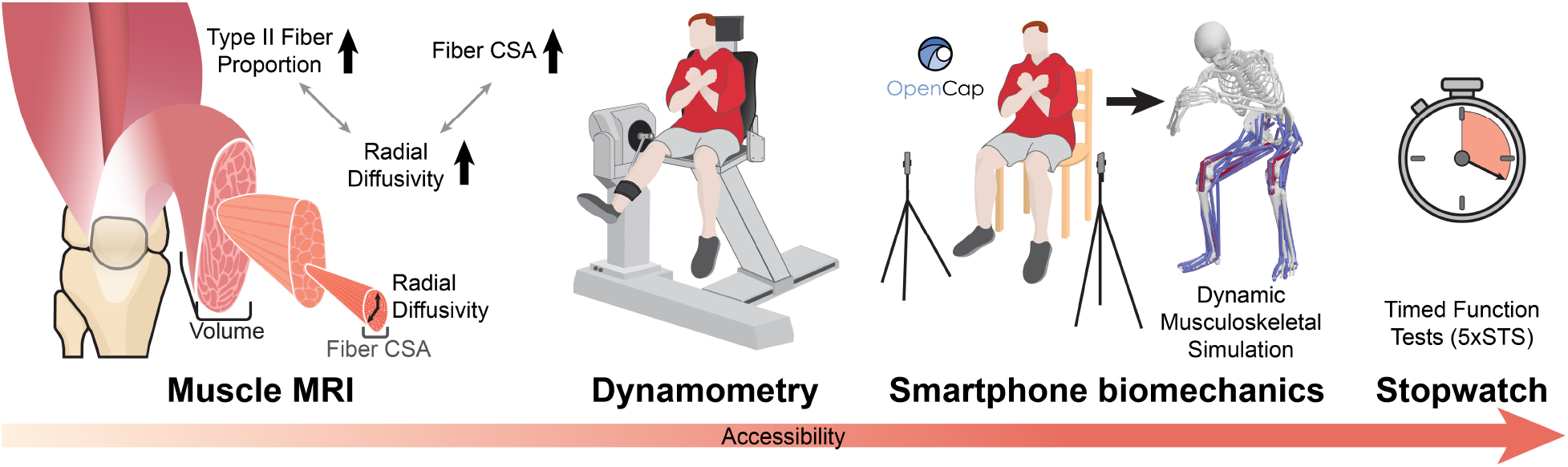
Four modalities used to measure quadriceps structure and function. MRI is a standard for quantifying muscle structure; we assess quadriceps volume and radial diffusivity, which relates to fiber cross sectional area (CSA) and type II fiber type proportion. Isokinetic dynamometry is the standard for assessing strength. The cost and time required for MRI and isokinetic dynamometry make them impractical for widespread assessments. Smartphone video–based biomechanical analysis could estimate both motion and joint moments in minutes, potentially offering a sensitive and accessible assessment of muscle function. Currently, timed function tests, such as the five times sit-to-stand (5xSTS) test, are commonly used in aging research since they only require a stopwatch, but they lack sensitivity to early decline in muscle function.

## Methods

Nineteen healthy individuals of various ages (57.8 ± 15.4 y; 30–78 y) participated in this study after consenting to a protocol approved by the Stanford University Institutional Review Board. Individuals performed all measurements (dynamometry and 5xSTS) prior to MRI during a single visit, except for one participant who returned two months after physical assessments for the MRI procedures due to technical issues with the scanner. Details of participant recruitment, exclusion criteria, and MRI/dynamometry protocols have been previously described [13]. Briefly, participants were healthy adults without contraindications to MRI or known neuromuscular or lower-extremity musculoskeletal conditions. Of the 24 participants previously described [13], 19 were included in the present analysis (Table 1) due to 5 having incomplete OpenCap data.

**Table 1:**
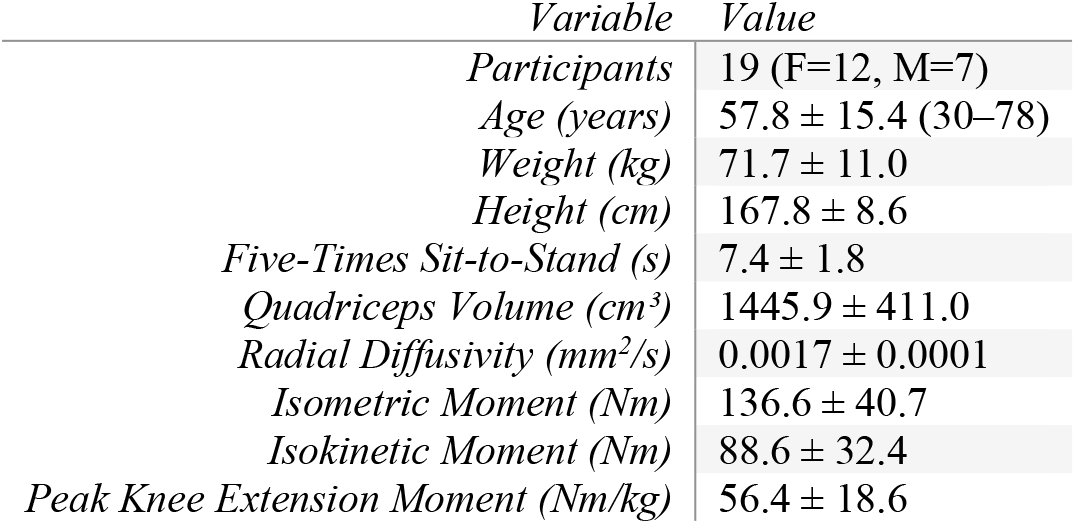
Participant demographics and key outcome measures for the analyzed cohort, reported as mean ± standard deviation.

| Variable | Value |
| --- | --- |
| Participants | 19 (F=12, M=7) |
| Age (years) | $57.8 \pm 15.4$ (30–78) |
| Weight (kg) | $71.7 \pm 11.0$ |
| Height (cm) | $167.8 \pm 8.6$ |
| Five-Times Sit-to-Stand (s) | $7.4 \pm 1.8$ |
| Quadriceps Volume ($\text{cm}^3$) | $1445.9 \pm 411.0$ |
| Radial Diffusivity ($\text{mm}^2/\text{s}$) | $0.0017 \pm 0.0001$ |
| Isometric Moment (Nm) | $136.6 \pm 40.7$ |
| Isokinetic Moment (Nm) | $88.6 \pm 32.4$ |
| Peak Knee Extension Moment (Nm/kg) | $56.4 \pm 18.6$ |

### MRI Acquisition and Processing

We computed volume and radial diffusivity in the vastus lateralis, vastus medialis, vastus intermedius, and rectus femoris muscles. Pulse sequence and processing details are described in Mazzoli et al., 2025. Briefly, MRI scans were performed on a 3T system (Signa Premier, GE Healthcare) using a 16-element anterior coil (AirCoil, GE Healthcare) on the thighs and the built-in 12 element posterior array. With participants in a supine position, we acquired Dixon multi-echo gradient-echo data for water/fat separation and muscle volume, and we used diffusion tensor imaging (DTI) for radial diffusivity. Quadriceps muscles were manually segmented by a trained rater, and diffusion data were non-rigidly registered to the Dixon scans. Lean muscle volume, defined as total volume multiplied by water fraction, was included in supplementary analyses to assess sensitivity to volume definition. To capture both muscle quantity and quality in a single feature, we standardized (i.e., created z-scores: zero mean, unit standard deviation) and summed total volume and radial diffusivity to form a composite MRI score of muscle quantity and quality as an exploratory metric. Similar to the individual features, increases in the composite metric should indicate increased force-generating capacity. Volume was not normalized to body mass, as the goal of this study was to examine associations in muscle force-generating capacity across measurement modalities rather than size-normalized performance. In this generally healthy cohort [13], body mass is mechanistically linked to muscle volume and strength, thus normalization would remove meaningful variance.

### Dynamometry

We measured knee extension moments using an isokinetic dynamometer (HUMAC NORM). Isometric moments were measured at 60°, and isokinetic moments were tested concentrically at 90°/s and 120°/s and eccentrically at 45°/s and 60°/s. We used isokinetic moments at 120°/s in our analysis since it is the most dynamic of the concentric movements collected. We provided participants standardized instructions that described the procedure and motivated maximal activity. For each condition, participants performed two submaximal practice contractions (~50% and ~75% effort) prior to 5 maximal contractions. We recorded the peak knee extension moment across maximal contractions for each leg. Moment recordings were corrected for gravity.

### Sit-to-Stand Assessment and Video-Based Biomechanics

Participants performed a maximal-effort 5xSTS test from a standardized chair height. A trained assessor recorded the 5xSTS time to completion using a stopwatch. Concurrently, we collected markerless motion capture with a two-smartphone OpenCap setup [33]. OpenCap automatically computed kinematics of an OpenSim model with 33 degrees of freedom and 80 muscles [37], [38], [39], and we performed dynamic muscle-driven simulations to estimate the knee extension moment [33]. We developed custom algorithms (Python, v. 3.12.2) to extract kinematic and kinetic parameters of interest from the kinematics and simulations. We analyzed the trunk orientation at lift-off (°), peak torso angular velocity prior to lift-off (°/s), and peak knee extension moment, averaged over the first three chair-rise repetitions from the 5xSTS test. These kinematic features were previously reported to compensate for quadricep weakness [28], and the knee extension moment has been related to quadriceps strength [34]. A prior analysis showed that this cohort had a high degree of symmetry in knee extension moments between left and right legs [13], so we used right leg MRI, dynamometry, and knee extension moment in the present analysis.

### Statistical Analysis

Associations between standardized MRI features (total volume, radial diffusivity, and the composite MRI score) and functional outcomes were assessed using univariate linear regression and Pearson correlation coefficients. Functional outcomes included isometric moment, isokinetic concentric moment (120 °/s), peak knee extension moment derived from OpenCap, 5xSTS time, trunk orientation at lift-off, and peak trunk angular velocity. We corrected for multiple comparisons by controlling for the false discovery rate using the Benjamini–Hochberg procedure [40]. We report corrected p-values with α=0.05. We compared peak knee extension moment (OpenCap, isokinetics, and isometrics), 5xSTS time, and kinematic features (torso orientation and angular velocity) with MRI parameters, and corrected for all p-value correlations in one group (18 tests).

## Results

OpenCap-derived peak knee extension moment during the 5xSTS correlated with quadriceps volume (r=0.63, p=0.014), radial diffusivity (r=0.61, p=0.016), and most strongly with the composite MRI score (r=0.77, p=0.002; Fig. 2).

**Fig. 2:**
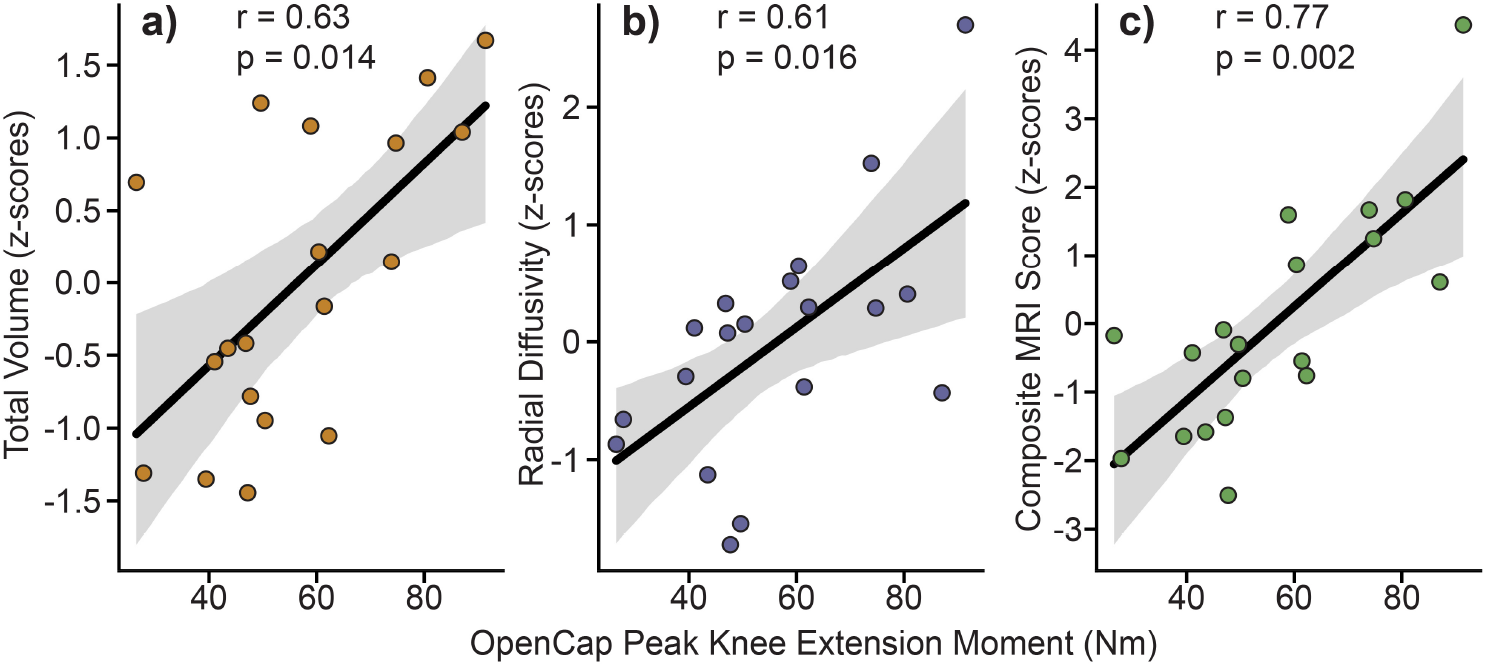
OpenCap-derived peak knee extension moment during five-time-sit-to-stand related to MRI metrics of **a)** total muscle volume, **b)** radial diffusivity, and **c)** a composite MRI score (volume + radial diffusivity). MRI metrics are shown as standardized z-scores. Lines of best fit (black) with 95% confidence intervals (gray) are shown.

The peak isokinetic knee extension moment from dynamometry correlated with volume (r=0.66, p=0.009) and the composite MRI score (r=0.73, p=0.002), but not radial diffusivity (r=0.52, p=0.054). The peak isometric moment from dynamometry only correlated with volume (r=0.75, p=0.002), not radial diffusivity (r=0.04, p=0.912) or the composite MRI score (r=0.49, p=0.066; Fig. 3). Correlations persisted across concentric and eccentric isokinetic dynamometry at varying speeds (Supplemental Fig. 1). The correlations between volume and OpenCap knee moment, isometric dynamometry, and isokinetic dynamometry were similar when using total versus lean muscle volume (Supplemental Fig. 2), consistent with the relatively low fat fraction of this cohort [13].

**Fig. 3:**
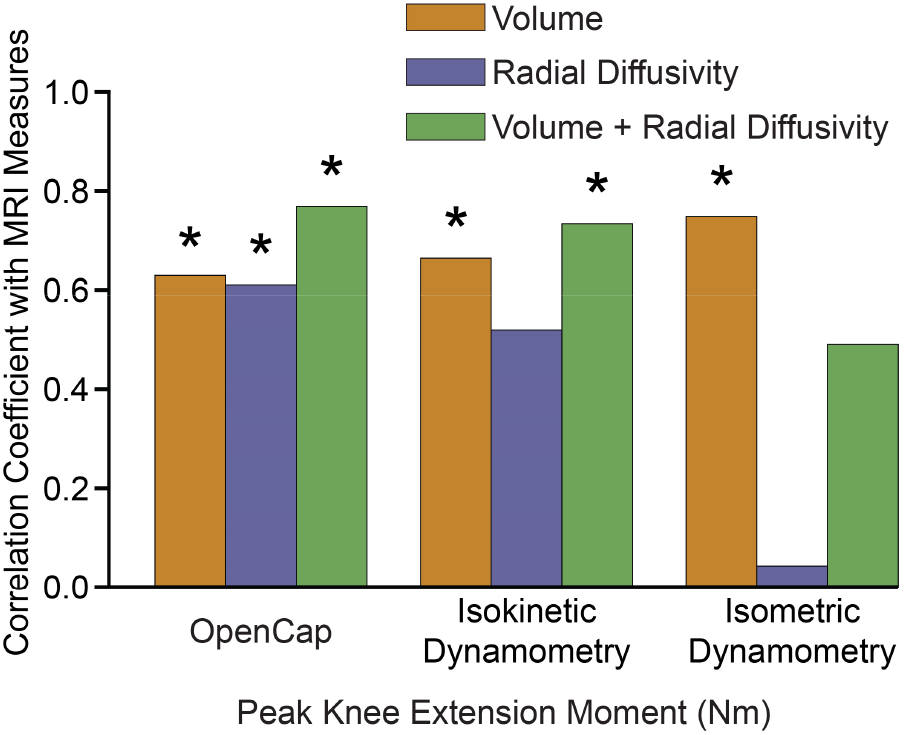
The peak knee extension moment from both OpenCap and concentric isokinetic dynamometry correlated with MRI-based measures of quadriceps force-generating capacity (volume and the composite MRI score). Isometric moments associated with volume, but not to radial diffusivity or the composite MRI score. *p<0.05

The 5xSTS time was not correlated with volume (r=0.01, p=0.973), radial diffusivity (r=−0.16, p=0.721), or the composite MRI score (r=−0.09, p=0.910). Torso orientation at liftoff was not associated with volume (r=0.35, p=0.223), radial diffusivity (r=0.05, p=0.913), or the composite MRI score (r=0.25, p=0.458), and peak torso angular velocity also did not significantly correlate with volume (r=0.51, p=0.054), radial diffusivity (r=0.06, p=0.913), or the composite MRI score (r=0.35, p=0.228; Fig. 4).

**Fig. 4:**
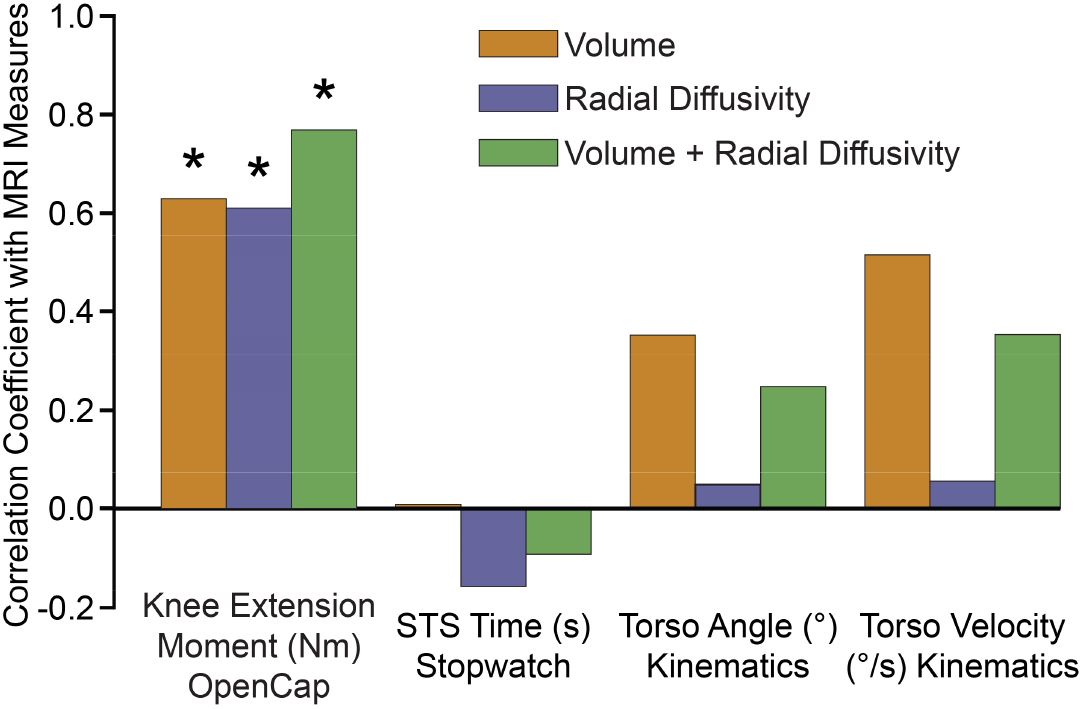
OpenCap-derived peak knee extension moment correlated with MRI metrics of quadriceps force-generating capacity (*p<0.05). 5xSTS time was not significantly correlated with any MRI metrics (p=0.721–0.973), nor were kinematic features (torso orientation and angular velocity; p=0.054–0.913).

## Discussion

The OpenCap-derived knee extension moment during chair rise was correlated with quadriceps volume, radial diffusivity, and the composite MRI score. Isokinetic moments from dynamometry showed largely similar strength correlations, but isometric dynamometry was only correlated with volume. The 5xSTS time, an accessible and commonly used clinical timed function test, was not correlated with any MRI metric. Because muscle strength depends on both quantity and quality of muscle, assessments incorporating dynamic force generation—whether through isokinetic dynamometry or OpenCap knee moment during maximal-effort chair rise—may better capture strength during functional tasks than isometric or spatiotemporal measures. Similarly, a composite score of volume and radial diffusivity produced the highest observed correlations with both isokinetic and OpenCap measured knee extension moments, indicating that both muscle volume and composition contribute to force-generating capacity during dynamic tasks. Importantly, OpenCap analysis of the sit-to-stand test can be conducted in less than five minutes, including equipment setup, using low-cost smartphones and free, open-source software. Augmenting the common 5xSTS test with OpenCap video-based kinetics provides a more informative, clinically accessible assessment of muscle function.

Muscle strength is a function of both quantity and quality [10], and it is strength—not muscle mass alone—that predicts mortality [41]. In our cohort, isometric moments associated with muscle volume but not radial diffusivity (a microstructural measure related to muscle quality) or the composite MRI score. These results align with prior findings. Isometric moments from dynamometry have been associated with muscle volume [7], [42], whereas previous studies have observed no association with radial diffusivity across healthy and aging populations [43], [44]. Instead, radial diffusivity has been linked to measures of dynamic force generation, including rate of force development [43] and isokinetic moment [45]. Collectively, this evidence suggests that isometric assessments are influenced by muscle volume but may not be sensitive to some microstructural measures of muscle quality, limiting their ability to fully capture muscle force-generating capacity during dynamic functional tasks.

The pattern of associations suggests that radial diffusivity is more strongly related to dynamic and force-velocity behaviors of muscle force production rather than isometric strength. Higher radial diffusivity has been related to larger fiber size, a greater proportion of type II (fast-twitch) fibers, and a faster rate of force development [15], [16], [43]. Thus, a muscle with higher radial diffusivity would be expected to generate greater force during high-velocity contractions. Radial diffusivity was associated with OpenCap-derived knee extension moment during 5xSTS, a functional task involving high-velocity contractions, but not with isometric moments. Although the association between radial diffusivity and isokinetic moments did not remain significant after false discovery rate correction, the strongest associations for both isokinetic- and OpenCap-derived knee extension moments were with the composite MRI score. Together, these results indicate that dynamic functional assessments capture features of muscle quantity and quality that independently contribute to muscle force-generating capacity. Since real-world mobility relies on mostly non-isometric muscle contractions, these dynamic functional assessments provide a more meaningful assessment of functional muscle performance than isometric tests. Similar patterns have been reported in other functional tasks, where knee moments during maximal-height jumping correlated with peak moments during high-velocity but not low-velocity isokinetic dynamometry [46]. Overall, these results demonstrate that the OpenCap-derived knee extension moment reflects both muscle volume and aspects of muscle quality, making it a useful indicator of force-generating capacity during functional activities.

While dynamometry remains the gold standard for assessing muscle strength, which relates to risk of falls and mortality [25], [41], it is not used in routine geriatric clinical care [20]. Instead, the 5xSTS is widely adopted in clinical practice due to its reliability [47] and feasibility [26]. Poor performance on the 5xSTS test indicates higher risk of disability and mortality [48], [49], and the test has been incorporated into National Institutes of Health and European consensus definitions of sarcopenia as a measure of physical performance [5], [50]. However, in our cohort of healthy adults from 30–78, we found no correlation between 5xSTS time and the MRI measures of muscle size or quality. This aligns with previous studies showing time-to-completion alone does not reflect muscle size, whether measured by MRI cross-sectional area [51] or DXA lean muscle mass [52]. It is possible that this relationship may be stronger in individuals with diagnosed sarcopenia, but the 5xSTS time is likely not sensitive to early changes in quadriceps force-generating capacity that precedes more dramatic declines in physical performance. Thus, while 5xSTS remains a valuable clinical tool, its time-to-completion is confounded by comorbidities and compensation strategies [53], [54], [55] which hinder its sensitivity to current muscle function [56].

Rising from a chair requires large quadriceps forces, and altered sagittal plane trunk kinematics can reduce quadriceps demand. Increased trunk flexion angle at seat-off moves the center of mass forward over the base of support, shifting demand from the knee extensors (quadriceps) to the hip extensors and ankle plantarflexors [28], [29], [35]. Greater trunk angular velocity prior to seat-off creates greater forward linear momentum that can be transferred into vertical momentum [36], also reducing knee extensor demand. We did not find correlations between trunk angle or peak angular velocity to the MRI metrics. This aligns with other studies that found no relationship between trunk angle and isometric strength [57], but conflicts with studies in older populations that have found relationships between angular velocity and grip strength [58]. Compared with our cohort, which spanned six decades, kinematic features may be more informative in older adults with frailty, who have been shown to use greater trunk range of motion during standing [59]. Our results highlight that a metric directly estimating peak quadriceps force-generating capacity—the knee extension moment during a quadriceps-demanding task—is more informative than surrogate kinematic measures or task-performance time. This further supports the potential utility of OpenCap-based *kinetic* assessments as a potential early indicator of declining muscle function.

It is important to identify the limitations of this study. First, our small cohort comprises healthy adults of various ages; however, validation in clinical populations and in longitudinal studies will be important. Prior work with this cohort found no differences in knee extension moments measured from dynamometry between legs [13], so pathologies with asymmetry warrant particular attention. We focused our analysis on the quadriceps given its role in fall risk and importance in ability to rise from a chair, which is a common and challenging activity of daily living.

Other muscle groups also contribute to sit-to-stand mechanics and other important activities of daily living [24]. For example, plantarflexor strength is important for stability and gait, and it declines with age [60], [61], [62]. Video-based kinetic assessments of other tasks that challenge other muscle groups could provide additional value in combination with the 5xSTS. Finally, while participants were instructed to perform the 5xSTS as quickly as possible, we did not use electromyography (EMG) to confirm maximal effort or muscle excitation. Any sub-maximal effort would bias the results towards weaker associations, meaning the correlations we observed are, if anything, conservative rather than inflated.

## Conclusion

MRI and isokinetic dynamometry remain the gold standards for assessing muscle quantity, quality, and force-generating capacity; however, neither are regularly used clinically. In contrast, the 5xSTS time is accessible but provided little insight into muscle force-generating capacity in our cohort of healthy participants of various ages. We showed OpenCap-derived knee extension moment during the 5xSTS related to both quadriceps volume and radial diffusivity, mirroring the relationships seen with isokinetic dynamometry. By capturing markers of muscle quantity and quality with a five-minute smartphone video–based assessment, OpenCap offers a promising approach to assessing lower-extremity muscle function that is more accessible than dynamometry or MRI, and more informative than the 5xSTS time alone. Furthermore, smartphone video–based assessments enable repeatable extraction of 5xSTS time and kinematic features, which in our unimpaired cohort were not associated with MRI-based measures of muscle function, but may become informative in populations with limited muscle function. This capability could substantially expand opportunities for routine, meaningful evaluation of muscle function in clinical care and research.

## Data Availability

All data presented here is from the open-source OpenCap platform that is freely available for academic research. All software and algorithms in this paper are similarly made open source and freely available (github.com/rdmagruder/mri-aging).

## Ethical standards

This study was approved by the Stanford University Institutional Review Board, and written informed consent was obtained from all participants prior to participation. All procedures were performed in accordance with the ethical standards of the institutional research committee and with the 1964 Declaration of Helsinki and its later amendments.

## Supplementary Material

We conducted exploratory analyses to determine how the chosen isokinetic motion and MRI-measures effected our results. In particular, we compared peak knee extension moments during eccentric, isometric, and concentric contractions to MRI-derived total volume, radial diffusivity, and composite score (Supplementary Fig. 1). The correlation strengths were higher with the faster movement, regardless of being eccentric or concentric. Also, we associated lean muscle volume (defined as total volume multiplied by water fraction) to OpenCap-derived knee extension moment and the peak knee moment measured by isometric and isokinetic dynamometry (Supplementary Fig. 2). The strengths of these correlations were similar to total muscle volume. These additional analyses were conducted to support the robustness, physiological plausibility, and methodological choices underlying the main results.

**Supplementary Fig. 1:**
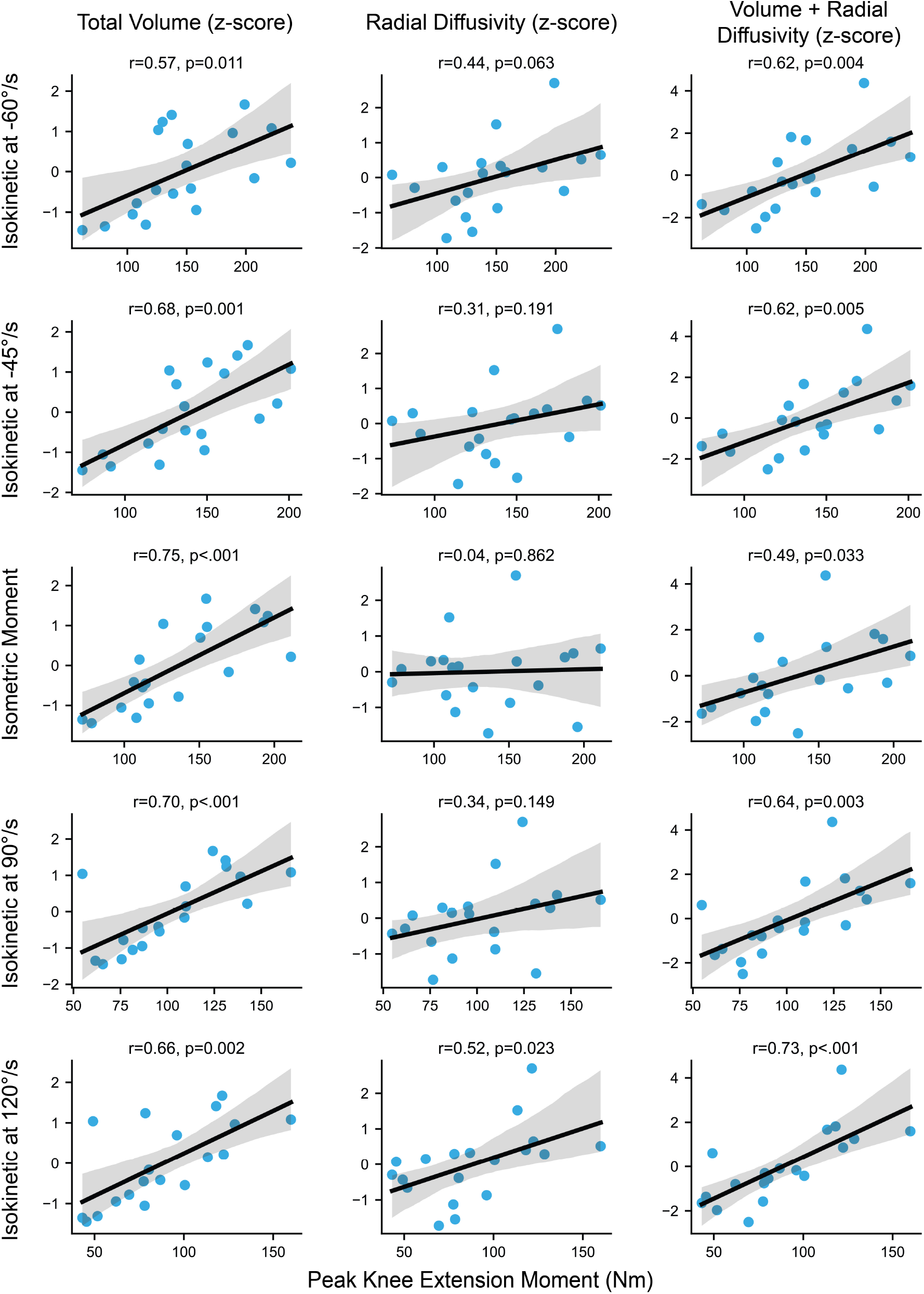
Peak knee extension moments from eccentric, isometric, and concentric contractions measured with dynamometry, correlated with MRI measures of volume and the composite MRI score. Higher velocities, both eccentrically and concentrically, correlated more strongly to radial diffusivity. Isokinetic moments at 120°/s were chosen since these contractions are maximally dynamic and are concentric, similar to the chair rise motion analyzed with OpenCap. Since this is an exploratory analysis, p-values are not corrected for multiple comparisons.

**Supplemental Fig. 2:**
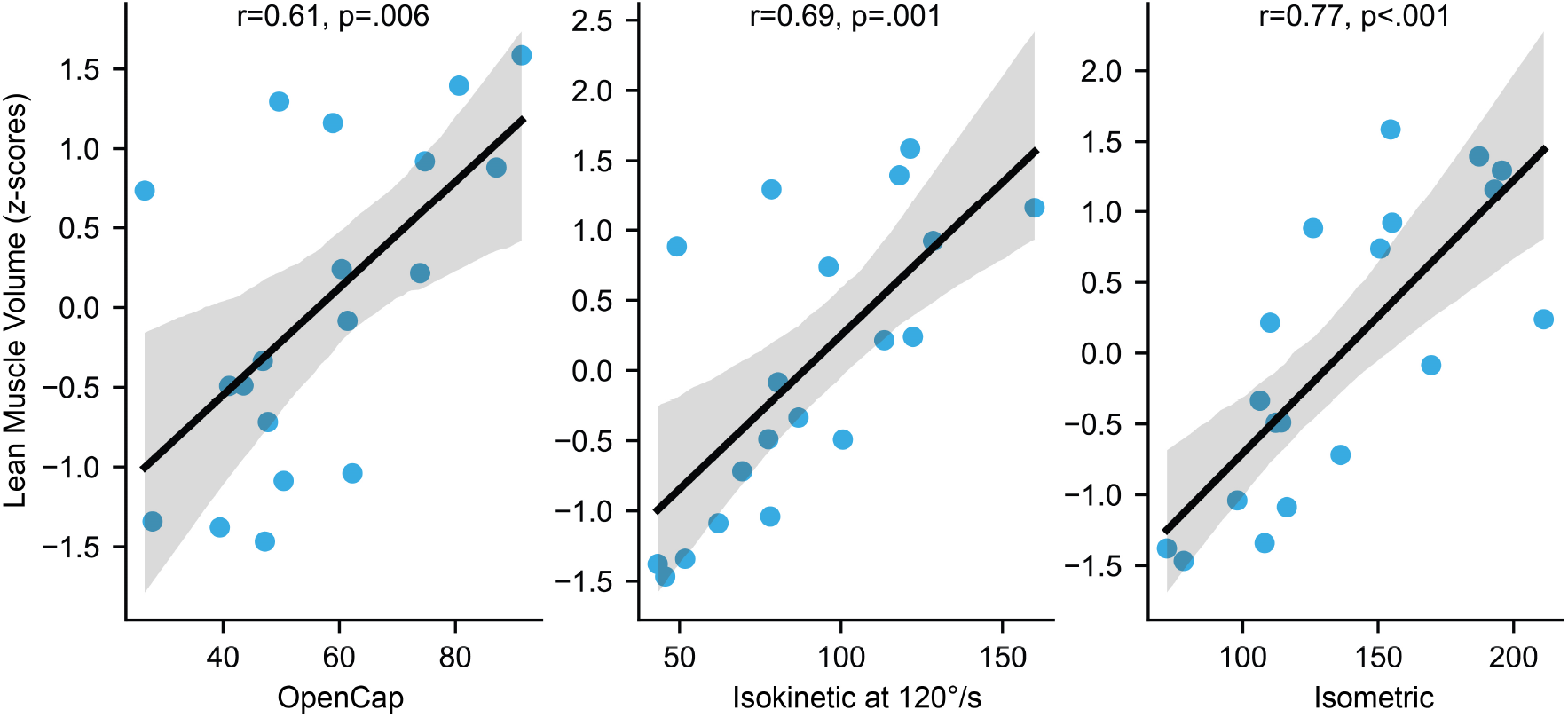
The peak knee extension moment measured with OpenCap, isokinetic dynamometry, and isometric dynamometry, correlated with MRI-measured lean muscle volume (r=0.61–0.77, p<0.01; not corrected for multiple comparisons). These associations are similar to those reported for total volume (r=0.63–0.75, p=0.002– 0.014; corrected for 18 tests).

